# SARS-CoV-2 infection-induced immunity and the duration of viral shedding: results from a Nicaraguan household cohort study

**DOI:** 10.1101/2022.06.17.22276565

**Authors:** Hannah E. Maier, Miguel Plazaola, Roger Lopez, Nery Sanchez, Saira Saborio, Sergio Ojeda, Carlos Barilla, Guillermina Kuan, Angel Balmaseda, Aubree Gordon

**Affiliations:** Department of Epidemiology, School of Public Health, University of Michigan in Ann Arbor, Michigan, USA; Sustainable Sciences Institute, Managua, Nicaragua; Centro Nacional de Diagnóstico y Referencia at the Ministry of Health, Managua, Nicaragua; Centro de Salud Sócrates Flores Vivas at the Ministry of Health, Managua, Nicaragua; Mount Sinai; Division of Infectious Diseases and Vaccinology, School of Public Health, University of California, Berkeley, California, USA

## Abstract

**Background:** Much of the world’s population has been infected with SARS-CoV-2. Thus, infection-induced immunity will play a critical role in future SARS-CoV-2 transmission. We investigated the impact of immunity from prior infection on viral shedding duration and viral load.

**Methods:** We conducted a household cohort study in Managua, Nicaragua with an embedded transmission study that closely monitors participants regardless of symptom status. Real-time reverse-transcription polymerase chain reaction (RT-PCR) and enzyme-linked immunosorbent assays (ELISAs) were used to measure infections and seropositivity, respectively. Blood samples were collected in Feb/March and Oct/Nov 2020 and 2021, and surrounding household intensive monitoring periods. We used accelerated failure time models to compare shedding times. Participants vaccinated ≥14 days prior to infection were excluded from primary analyses.

**Results:** There were 600 RT-PCR-confirmed SARS-CoV-2 infections between May 1, 2020 and March 10, 2022 with ELISA data prior to infection. Prior infection was associated with 48% shorter shedding times, event time ratio (ETR) 0.52 (95% CI: 0.39-0.69, mean shedding: 13.7 vs 26.4 days). A 4-fold higher anti-SARS-CoV-2 spike titer was associated with 17% shorter shedding (ETR 0.83, 95% CI: 0.78-0.90). Similarly, maximum viral loads (lowest CT) were lower for previously infected individuals (mean CT 29.8 vs 28.0, p = 4.02×10^−3^). Shedding was shorter in previously infected adults and children ≥10 years, but not in children 0-9 years; there was little difference in CT levels for previously infected vs naïve adults above age 60.

**Conclusions:** Prior infection-induced immunity was associated with shorter viral shedding and lower viral loads.

---

As the SARS-CoV-2 pandemic continues into its third year, over 40% of the world’s population has been infected (Covid-19 Cumulative Infection Collaborators, 2022), making it critical to understand how immunity from prior infection affects repeat infection and transmission, particularly for low- and middle-income countries where vaccination rates are lower (Ritchie et al., 2020).

We used data from an existing household cohort study of 2,539 individuals 0 to 94 years of age in Managua, Nicaragua (Maier, Kuan, et al., 2021). We compare the duration of SARS-CoV-2 viral shedding and viral load between previously infected and serologically naïve individuals and examine the association of antibody levels with viral shedding duration.

## METHODS

### The Household Influenza Cohort Study (HICS)

HICS is an ongoing prospective cohort study of influenza in households that are free of disease at baseline (Figure S1). Located in district II of Managua, Nicaragua, HICS began in 2017 and was expanded in February 2020 to examine SARS-CoV-2 infection and disease. At the first indication of any illness, participants are requested to report to the study health center, where they are provided with their primary care. A transmission study is nested within HICS (Figure S2), in which participants are monitored closely and tested regardless of symptoms once a SARS-CoV-2 case is detected in their household allowing for the detection of cases regardless of symptoms. Once households are activated, study staff visit regularly, at approximately days 0, 3, 7, 14, 21, and 30, to collect combined nasal/oropharyngeal swabs and symptom diaries. Blood samples for serology were collected annually in March-April (annual samples) and October-December (midyear samples) and surrounding intensive monitoring periods (at household activation and 30-45 days later).

This study was approved by the institutional review boards at the Nicaraguan Ministry of Health and the University of Michigan (HUM00119145 and HUM00178355). Informed consent or parental permission was obtained for all participants. Assent was obtained from children aged ≥6 years.

### Laboratory Assays

Real-time reverse-transcription polymerase chain reaction (RT-PCR) was performed according to the protocol from Chu et al. (Chu et al., 2020). Enzyme-linked immunosorbent assays (ELISAs) were run on paired serum samples (current vs baseline) with a protocol adapted from the Krammer laboratory (Amanat et al., 2020). The SARS-CoV-2 spike receptor binding domain (RBD) and spike proteins for ELISAs were produced in single batches at the Life Sciences Institute at the University of Michigan. For ELISAs, RBD was used for screening (positive/negative) and spike was used for titers. The limits of detection for endpoint titers were 100 (lower) and 6,400 (upper). RBD screens and spike titers were run on 2020 annual and midyear and 2021 annual samples and for seronegative individuals on all samples; spike titers without RBD screens were run on 2021 midyear and intensive monitoring samples for individuals that were previously seropositive. All RT-PCR and most ELISAs were performed at the Nicaraguan National Virology Laboratory, with a minority of 2020 annual samples processed at the University of Michigan. Sequencing information was described previously (Maier, Balmaseda, et al., 2021).

### Viral shedding

RT-PCR-positive episodes were considered separate episodes if they were ≥60 days apart. Viral shedding durations were defined as either 1) ≥ the number of days RT-PCR-positive (right censored) or 2) between the number of days RT-PCR-positive and the time between prior and subsequent negative RT-PCR tests (interval censored).

### Vaccination

To assess seropositivity resulting from prior infection, individuals with any vaccine dose ≥14 days prior to shedding onset were excluded for primary analyses. Additionally, an analysis was run to compare full vaccination to otherwise seropositive (could have incomplete vaccination) and seronegative.

### Analysis

Participant age was calculated at the time of infection. Antibody titers were log-transformed and rounded for all analyses (log4(titer/5)) to reflect serial dilutions (original titer values of 5, 80, 320, 1280, 5120 were analyzed as 0, 1, 2, 3, 4, 5). Detectable spike titers without an RBD screen were coded as seropositive and negative RBD-screened samples without a spike titer were coded as a non-detectable spike titer. Censored shedding times were stored as survival objects (e.g. 1+, 15+ or [1,6], etc days) using the survival package (Thernau, 2021), Figure S3. Accelerated failure time models were used to compare the shedding survival objects by prior immunity, using the ‘survival’ (survreg function with a Weibull distribution) and ‘SurvRegCensCov’ (weibullReg function) packages (Hubeaux & Rufibach, 2015; Thernau, 2021). To compare viral load by prior immunity, we used Wilcoxon rank-sum tests (serostatus), linear regression (titer), and plotted by age with loess smoother (ggplot’s geom_smooth). All analyses were performed in R version 4.1.2. (R Core Team, 2021) using the tidyverse (Wickham et al., 2019) and code is available (Maier, 2022).

## RESULTS

Three SARS-CoV-2 waves occurred in Managua, taking place roughly from May-July 2020, April-October 2021, and January-March 2022 (Figure 1A). The second wave was predominantly gamma and delta variants (Maier, Balmaseda, et al., 2021), and third was presumably omicron (Hodcroft & Neher, 2022). Across the three waves, 745 RT-PCR confirmed SARS-CoV-2 infections were detected (Table S1, Figure S4&5); 262 (35%) of infections were in index cases and 483 (65%) were in household contacts. The full cohort of 2,539 participants aged 0 to 94 years with average household size of 5.1 people (range 2-12) has been described previously (Maier, Balmaseda, et al., 2021; Maier, Kuan, et al., 2021)). Blood samples for ELISAs nicely book-ended the first 2 waves and preceded the 3^rd^ wave (screening and titer samples preceding infections, and their results, are shown in Figure 1B-E). The overall mean shedding duration was 17.1 days (IQR: 9.91-26.3 days). There were 145 infections having ≥1 vaccine dose ≥14 days prior which were excluded from primary analysis, and 53 infections among those fully vaccinated ≥14 days prior (Table S1, Figure 1F).

**Figure 1.**
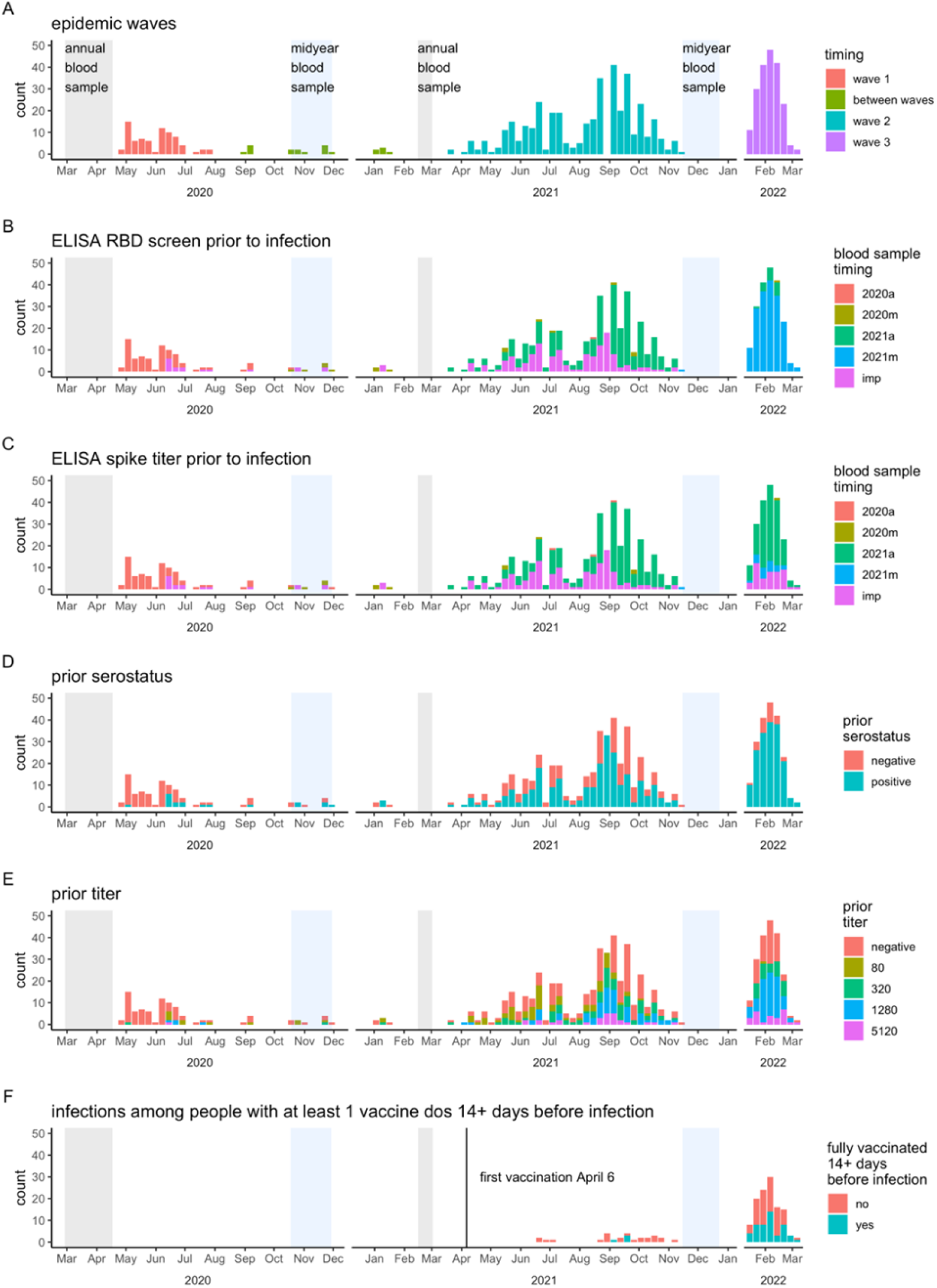
Epidemic timing. Weekly RT-PCR counts are colored according to A) epidemic wave, B-C) most recent blood sample for ELISA RBD screen and spike titer, D-E) RBD screen and spike titer results, and F) full vaccination status. Panel F) is subset to show only infections in people with ≥1 vaccination ≥14 days prior to infection.

### Prior infection and viral shedding duration

Overall, prior infection was associated with 48% shorter shedding (event time ratio [ETR]: 0.52, 95% CI: 0.39-0.69, Figure 2A). Mean shedding for prior infected versus naïve individuals were 13.7 days (IQR: 8.1-20.7) versus 26.4 days (IQR: 15.7-39.9, Figure 2A). A 4-fold higher spike titer was associated with 17% shorter shedding (ETR 0.83, 95% CI: 0.78-0.90, Figure 2B); those with the highest titers of 5120 shed on average 10.2 days (IQR 6.0-15.4). Full vaccination was associated with a similar level of shortened shedding (ETR fully vaccinated vs seronegative: 0.46, 95%CI: 0.31-0.68, Figure 2C).

**Figure 2.**
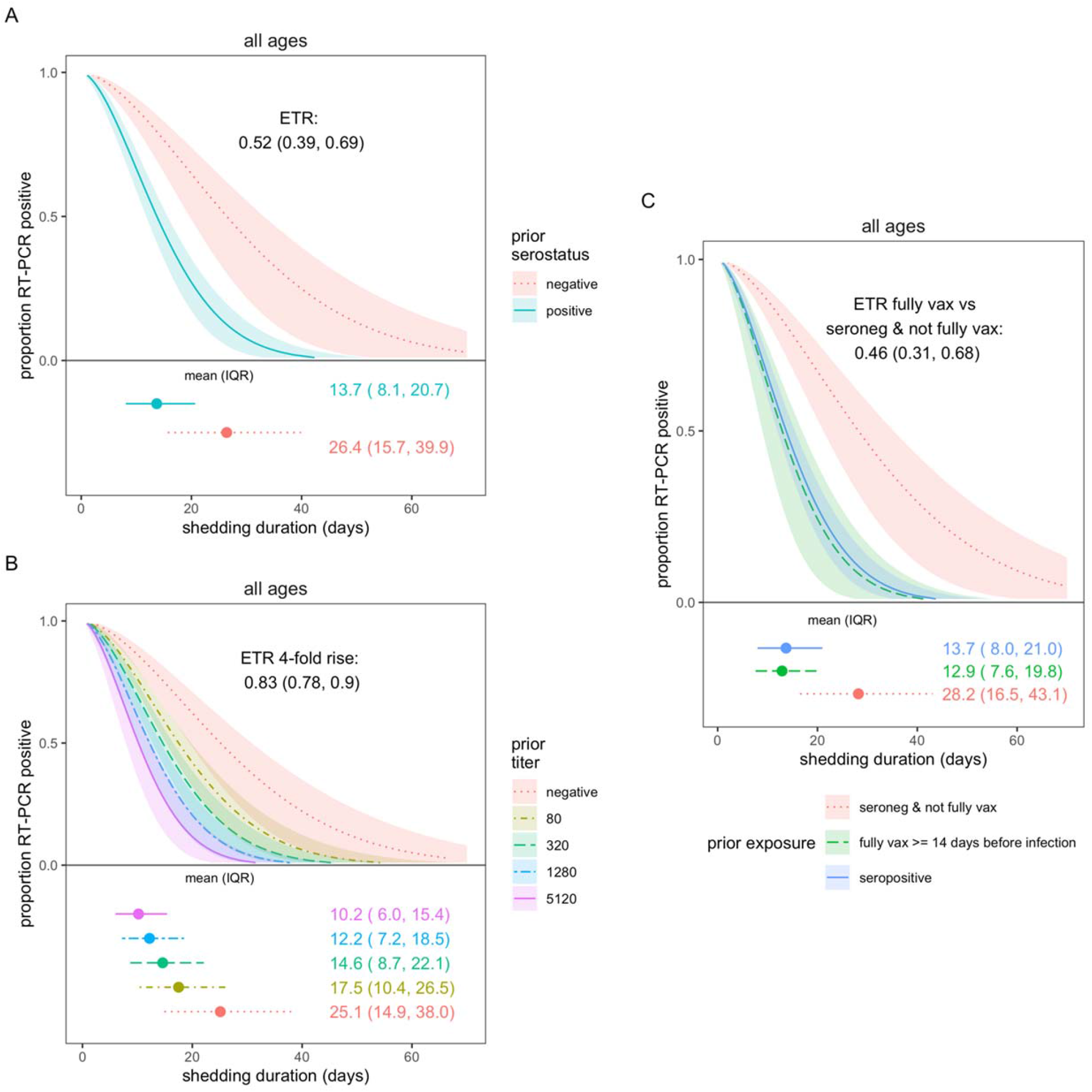
SARS-CoV-2 viral shedding duration by prior immunity. Prior immunity was measured as A) serostatus and B) anti-spike titer. C) compares fully vaccination and serostatus. Results are from accelerated failure time (AFT) models. Shaded regions represent 95% confidence intervals. Individuals with ≥1 vaccination ≥14 days prior to infection were excluded from analyses in A) and B).

In adults and older children (10-17 years), prior infection was associated with shortened shedding (ETRs: 0.31, 95%CI: 0.17-0.56 and 0.54, 95%CI: 0.32-0.88, respectively), but there was little difference for children 0-9 years by prior infection status (ETR 0.77, 95%CI: 0.46-1.28, Figure 3A). Naïve adults shed 3 times as long as naïve children aged 0-9y (ETR 3.1, 95%CI: 1.48-6.52) but there was no difference in shedding times between prior infected children and adults (Figure S6). Similarly, 4-fold higher titers in adults and older children were also associated with shorter shedding, but not significantly associated for children aged 0-9 years (Figure 3B, ETRs for adults, older children, and younger children: 0.78, 95%CI: 0.70-0.88; 0.84, 95%CI: 0.74-0.96; and 0.88, 95%CI: 0.76-1.02, respectively).

**Figure 3.**
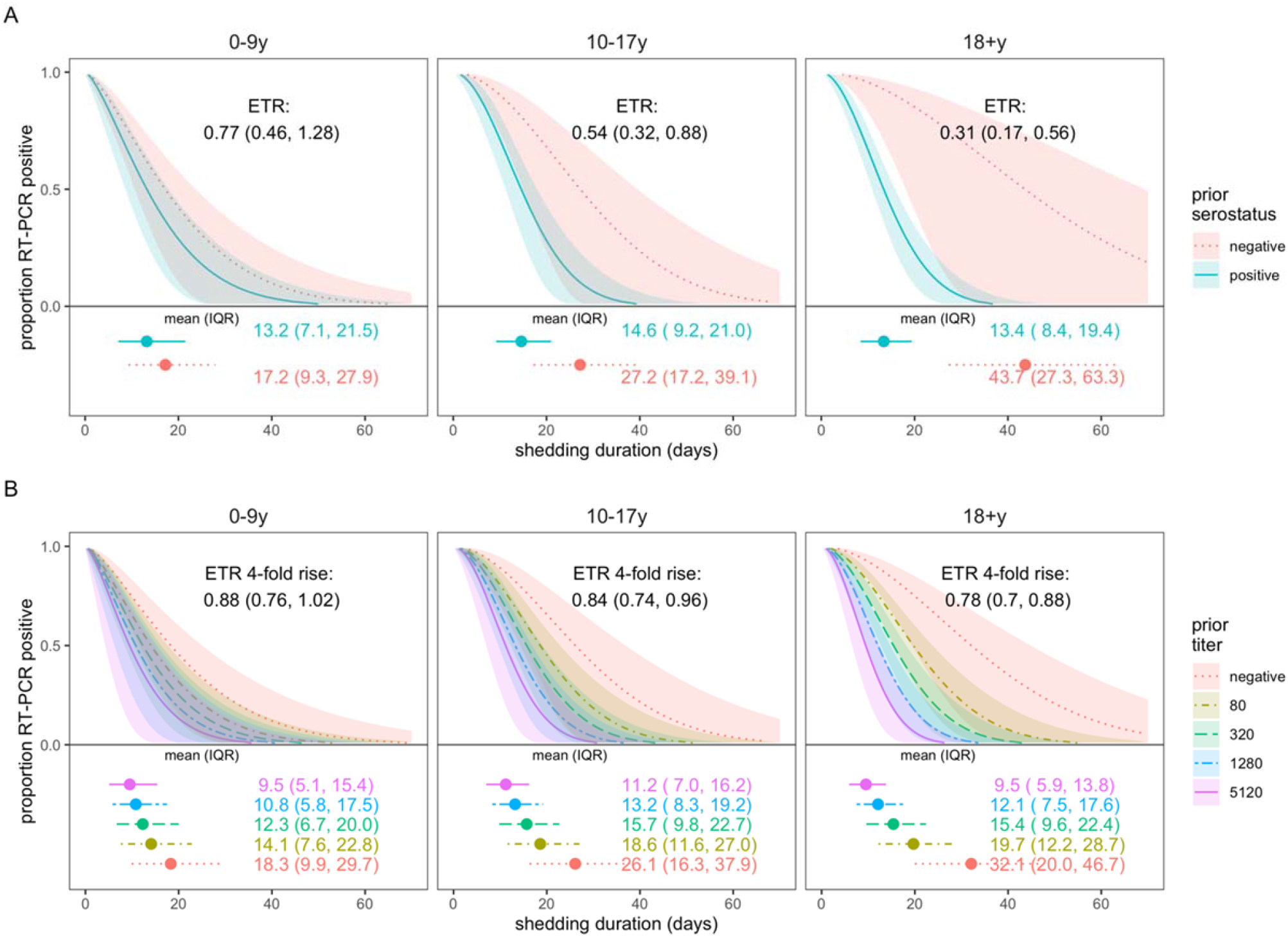
SARS-CoV-2 viral shedding duration by prior immunity and age. Prior immunity was measured as A) serostatus and B) anti-spike titer. Results are from accelerated failure time (AFT) models. Shaded regions represent 95% confidence intervals. Individuals with ≥1 vaccination ≥14 days prior to infection were excluded.

### Prior infection and viral load

Maximum viral loads, measured by RT-PCR cycle threshold (CT; higher CT = lower viral load), were slightly lower for previously infected vs naïve individuals (mean 29.8 vs 28.0 cycles, p = 0.0004,, Figure 4A). Higher anti-spike titers were also associated a lower maximum viral load (32.1 vs 28.3 cycles for titers of 5120 vs negative Figure 4C). Viral loads were somewhat lower across all ages for those with prior immunity (Figure 4B&D), except for participants above age 60y.

**Figure 4.**
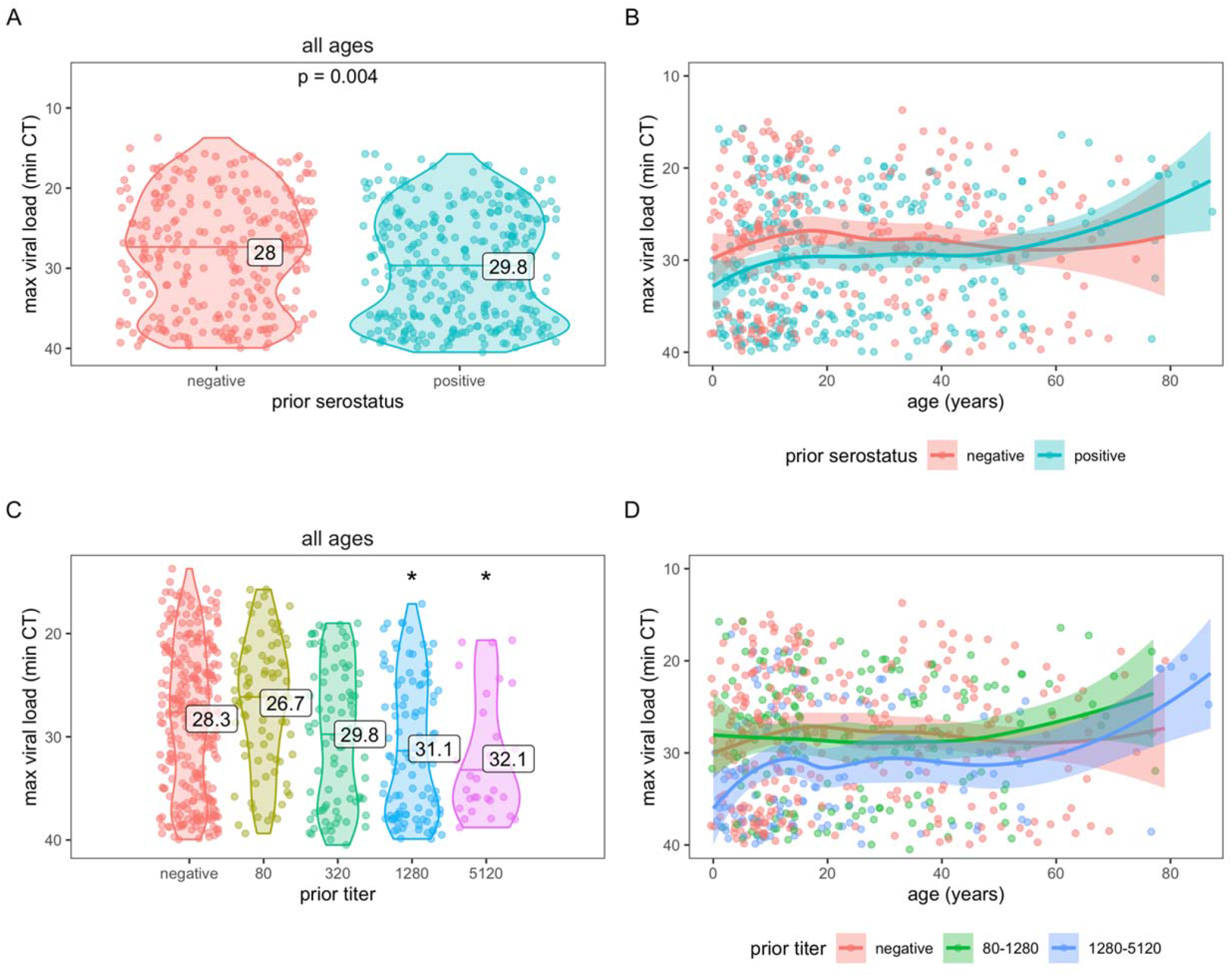
SARS-CoV-2 viral load by prior immunity and age. Prior immunity was measured as serostatus (A and B) and anti-spike titer (C and D). Each data point represents an infection. Violin plots (A and C) show the distribution of viral load, by each level of immunity; horizontal lines indicate median CT values and printed numbers represent mean CT values. Asterisks in C) indicate mean CT values significantly different from those in the negative group (p<0.05). Line plots (B and D) are fitted with a loess smoother and shaded regions represent 95% confidence intervals.

## DISCUSSION

We found that prior infection-induced immunity was associated with shorter shedding duration in adults and older children, but not in younger children. And adults shed longer than children among the naïve but similarly among previously infected. Although RT-PCR is not a direct indicator of viral viability or infectiousness, RT-PCR trajectories do roughly track with viable virus (Killingley et al., 2022), so shorter RT-PCR-detected shedding among prior infected will likely still translate to lower transmission. These results suggest that as immunity is established, children may contribute proportionally more to transmission, and transmission will decrease (due to both lower susceptibility and lower transmissibility from shorter shedding times).

Our long shedding times are in line with what others have found—a meta-analysis of 79 studies also found a mean shedding time of 17 days (Cevik et al., 2021). Vaccination has been shown to shorten viral shedding (Singanayagam et al., 2022). Higher viral loads (CT) have been found in reinfections (4.0 cycles) and vaccine breakthrough infections (1-3 cycles) (Abu-Raddad et al., 2022). We could not find any other studies comparing viral shedding duration by level of prior immunity.

A limitation of our work is that blood samples were not available shortly before all infections—a few were from ∼a year prior (Figure 1B&C). During long delays, 1) subsequent undetected infections could occur and 2) antibody titers may decline. Fortunately, the study design allows us to detect many even inapparent infections. And if the antibodies were lower than at time of measurement, we would expect to see an even stronger association. We did not have a large enough sample size (with the censored data) to look at viral shedding in older adults, as we did with viral load.

In addition to vaccination, prior infection will have a major impact on the future of the SARS-CoV-2 pandemic and should be duly considered.

## Data Availability

DATA AND CODE AVAILABILITY Individual-level data may be shared with outside investigators following University of Michigan IRB approval. R code is available on GitHub (https://github.com/hannahma/SARS-CoV-2_shedding). Please contact Aubree Gordon to arrange for data access.

## Funding

This work was supported by the National Institute for Allergy and Infectious Diseases at the National Institute of Health [award no. R01 AI120997 to A.G., and contract nos. HHSN272201400006C and 75N93021C00016 to A.G.], and a grant from Open Philanthropy.

## ACKNOWLEDGEMENTS

We are extremely grateful to the families who participated in this study and to the incredibly dedicated teams at the Centro de Salud Sócrates Flores Vivas, the Nicaraguan National Virology Laboratory at the Nicaraguan Ministry of Health and the Sustainable Sciences Institute who have continued to work through this pandemic. We would like to thank Leo Poon for providing the protocol and controls for RT-PCR testing, and Florian Krammer for sharing RBD and Spike constructs as well as technical advice. We are grateful to Janet Smith, Melanie Ohi and their groups at the Center of Structural Biology at the UM Life Sciences Institute for producing proteins and antibodies for the ELISAs.

## COMPETING INTERESTS

Aubree Gordon serves on an advisory board for Janssen and has received consulting fees from Gilead Sciences. All other authors report no competing interests.

## DATA AND CODE AVAILABILITY

De-identified data needed to create the figures and R code is available on GitHub (https://github.com/hannahma/SARS-CoV-2_shedding). As this is a human subjects study, the full data are not publicly available. However, individual-level data may be shared with outside investigators who submit a proposal for review by the study executive committee following University of Michigan and Nicaraguan IRB approval.”

## Supplement for

**Table S1.**
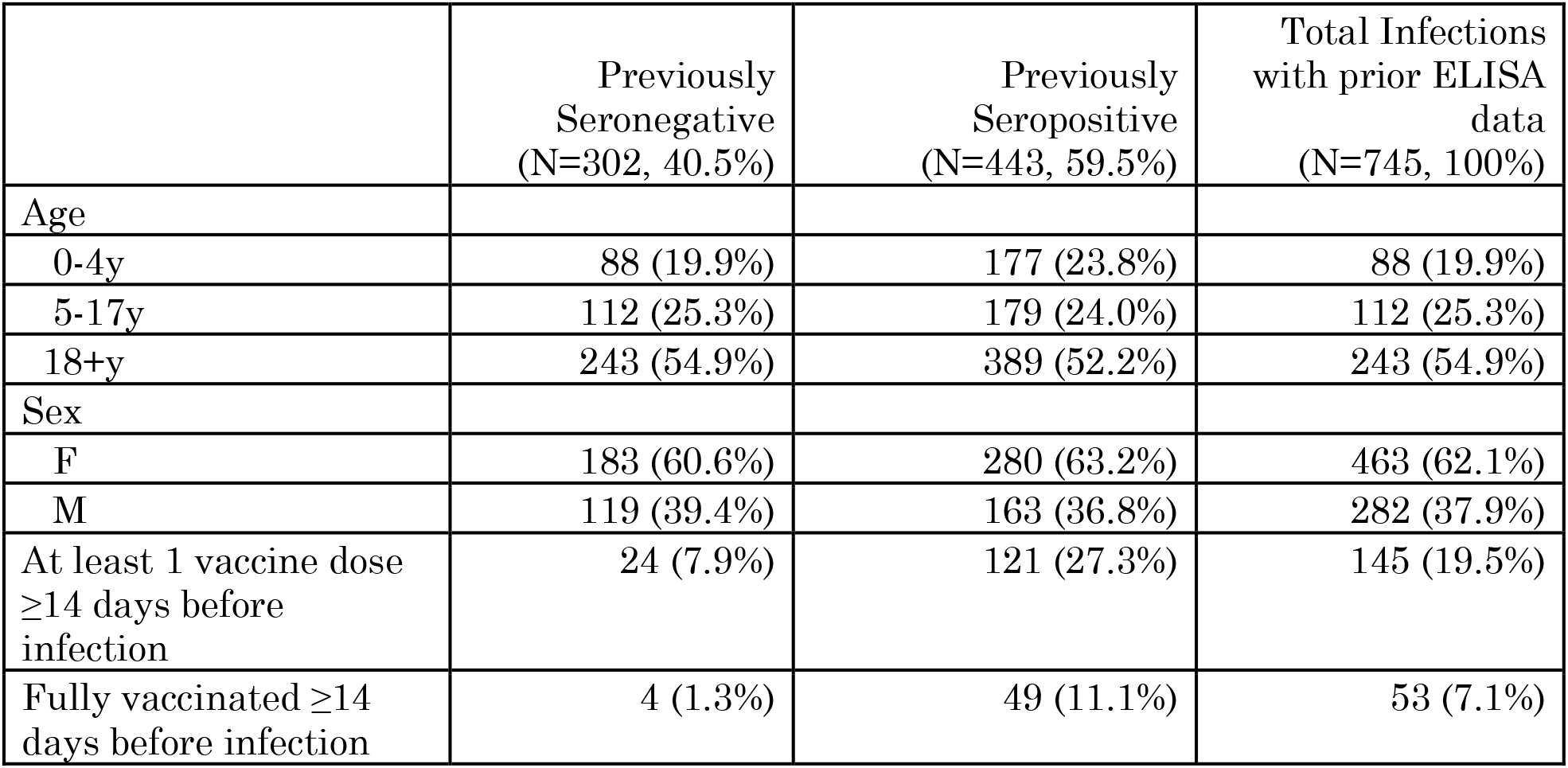
Characteristics of SARS-CoV-2 infections

**Figure S1.**
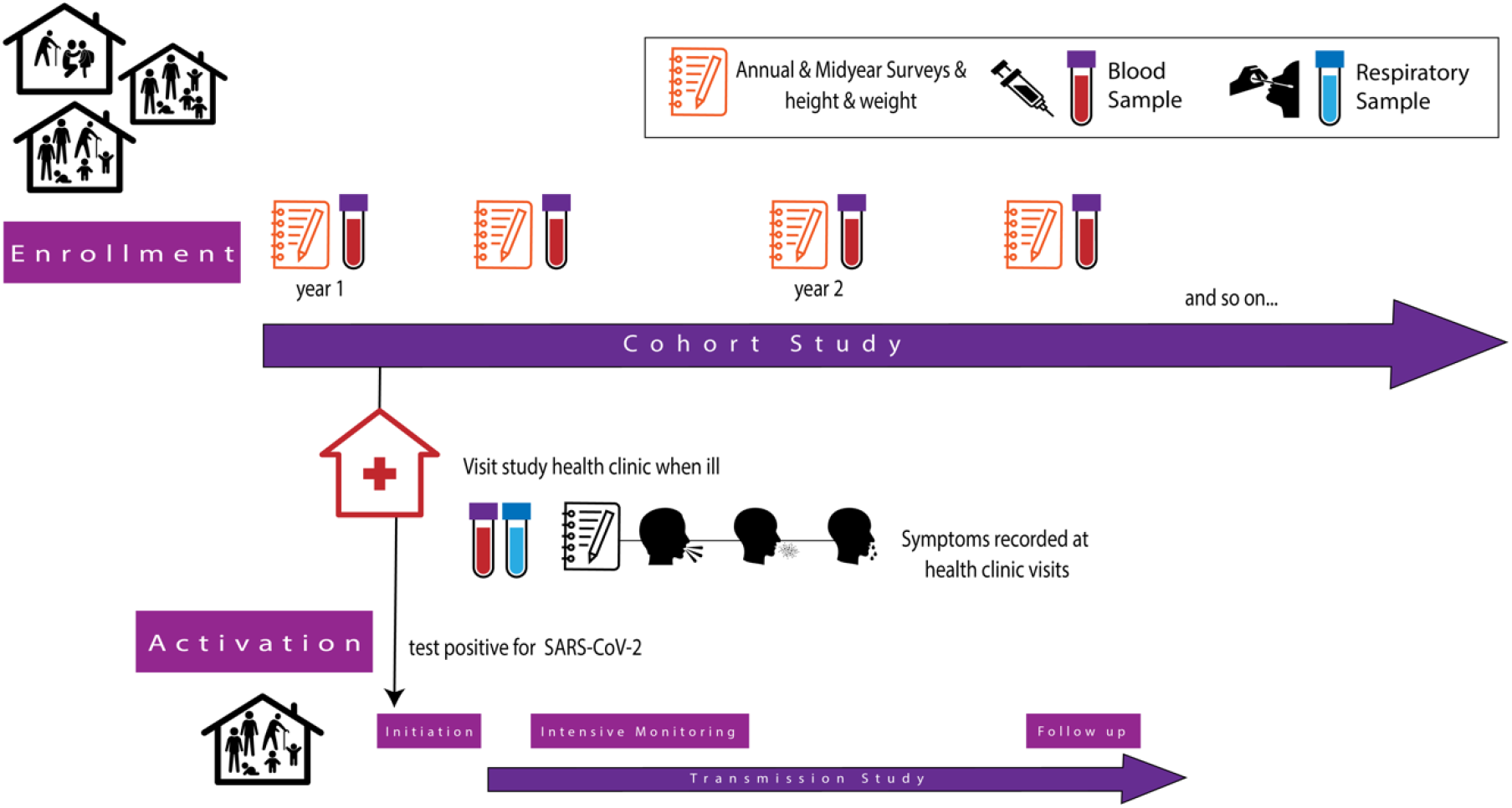
Cohort study diagram for HICS.

**Figure S2.**
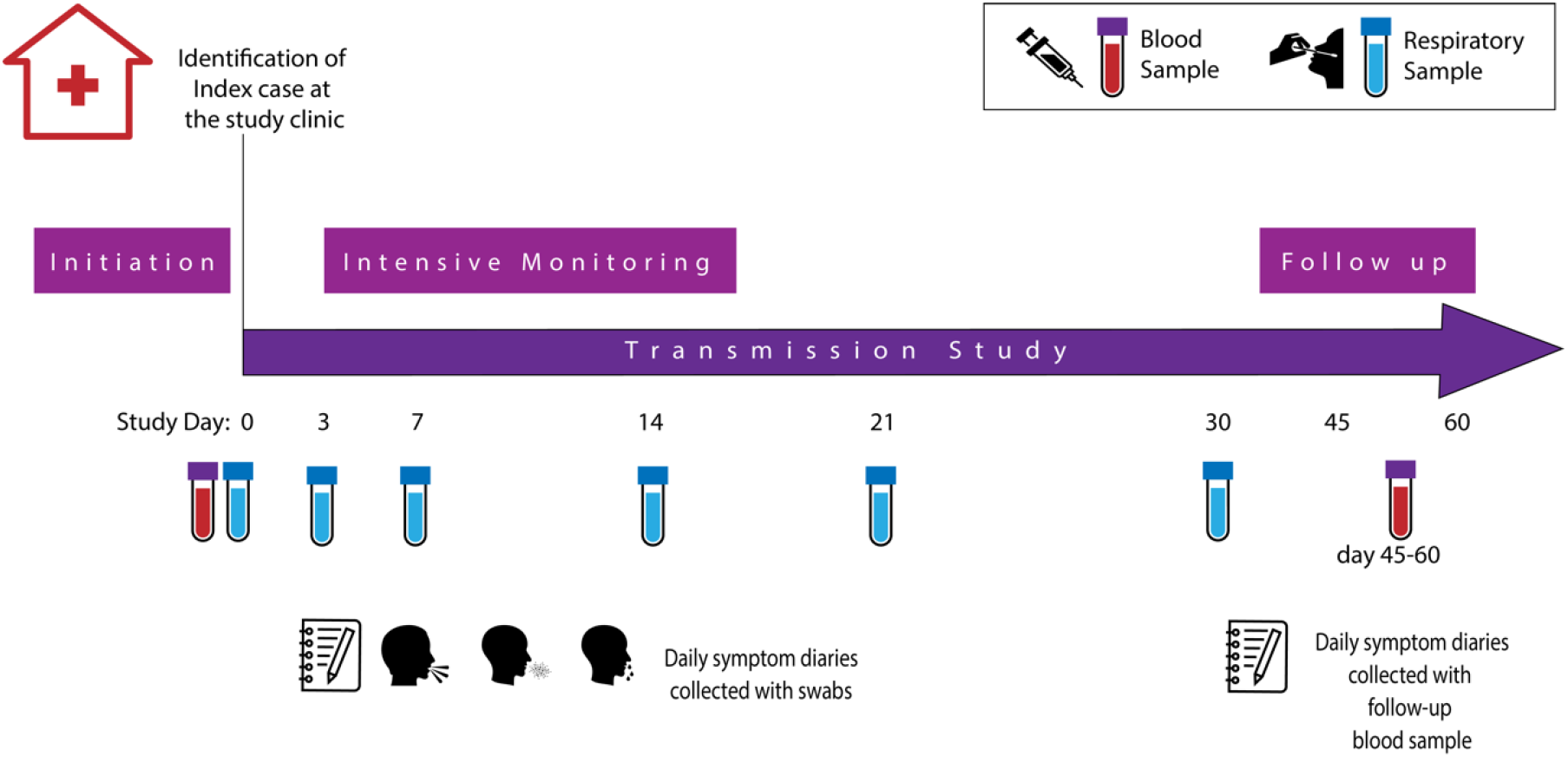
Nested transmission study diagram for SARS-CoV-2 activations in HICS.

**Figure S3.**
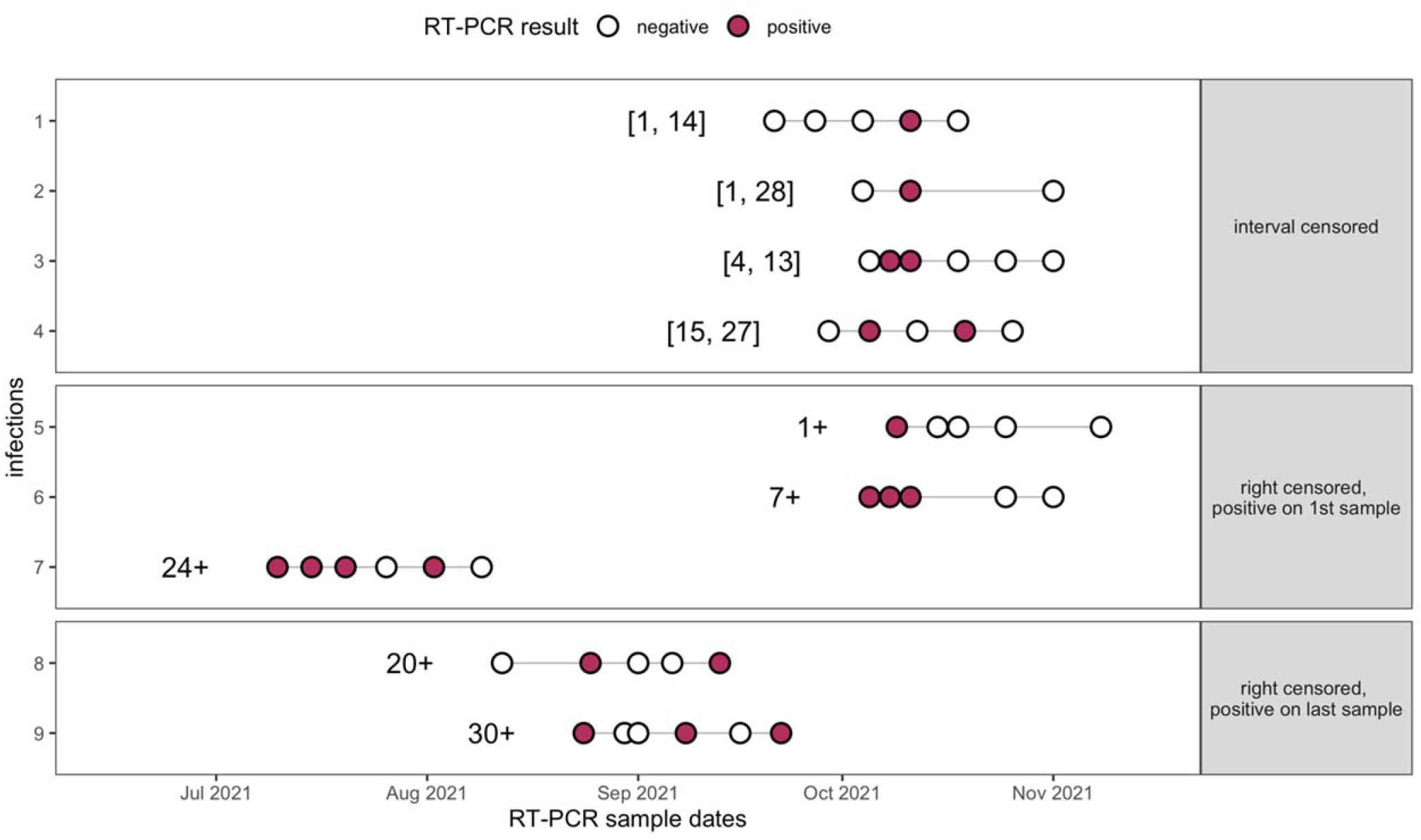
Example of shedding time censoring. Shedding times listed are formatted as survival objects, created with the ‘survival’ package.

**Figure S4.**
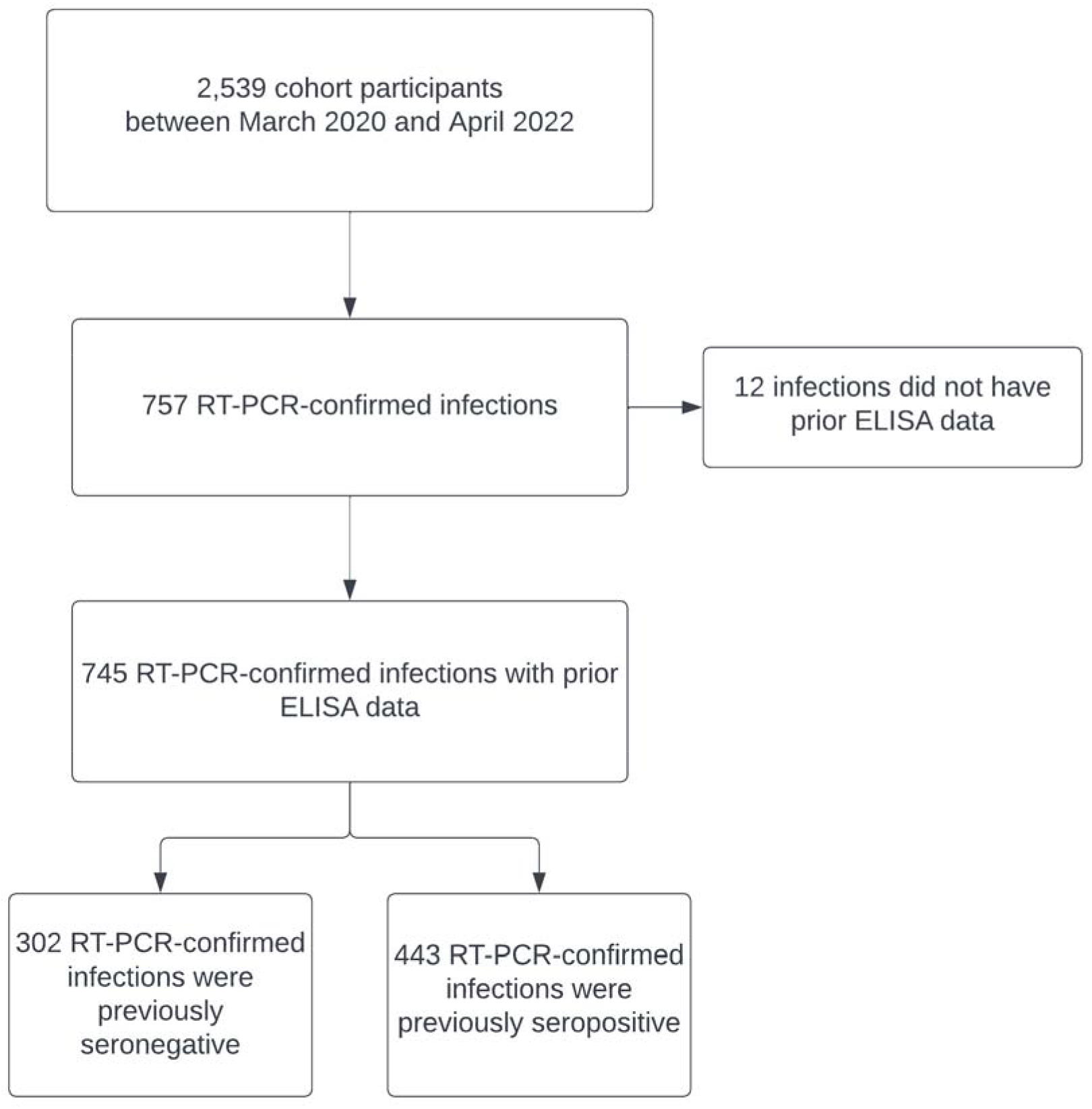
Study flowchart.

**Figure S5.**
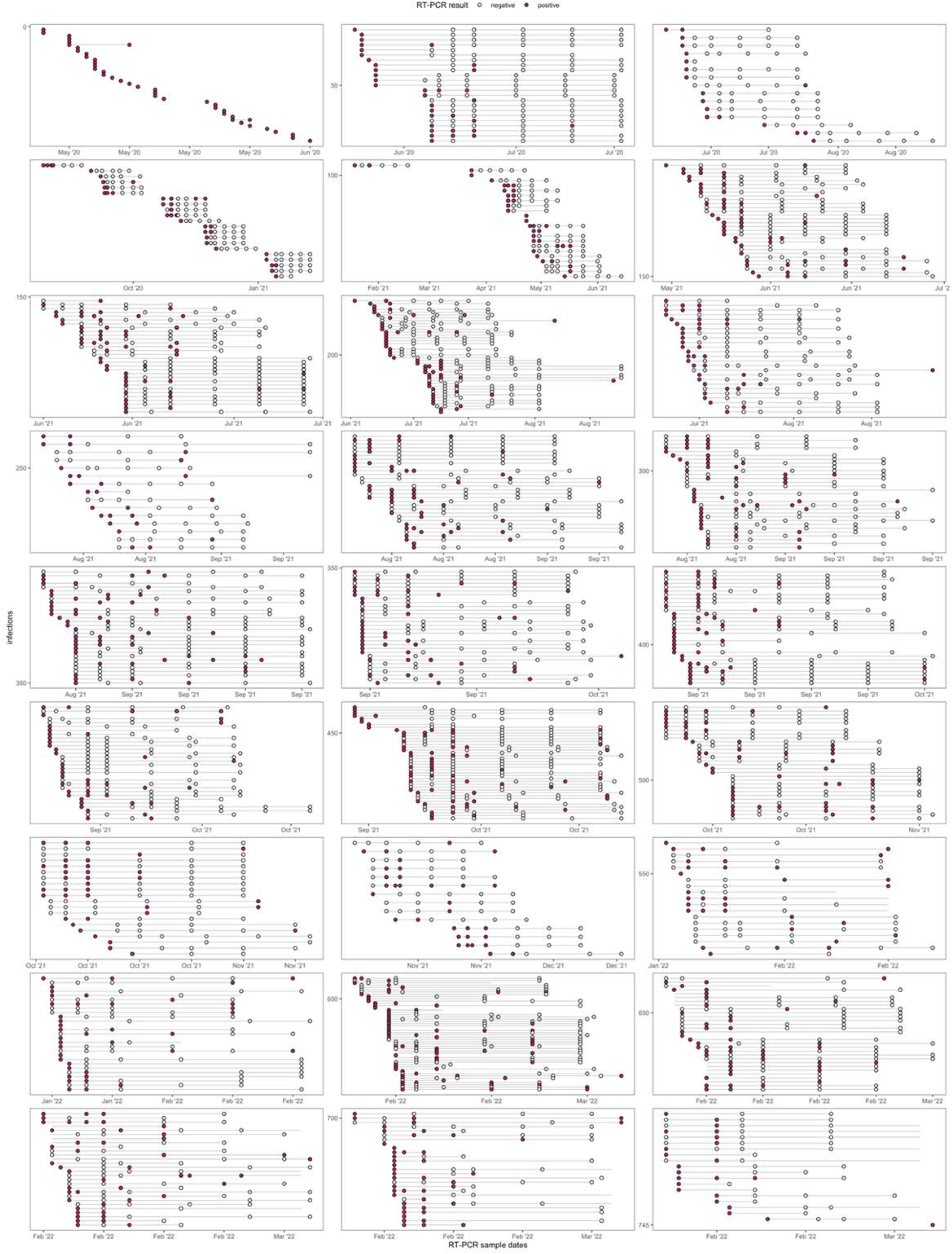
Sampling and RT-PCR results for all 745 infections.

**Figure S6.**
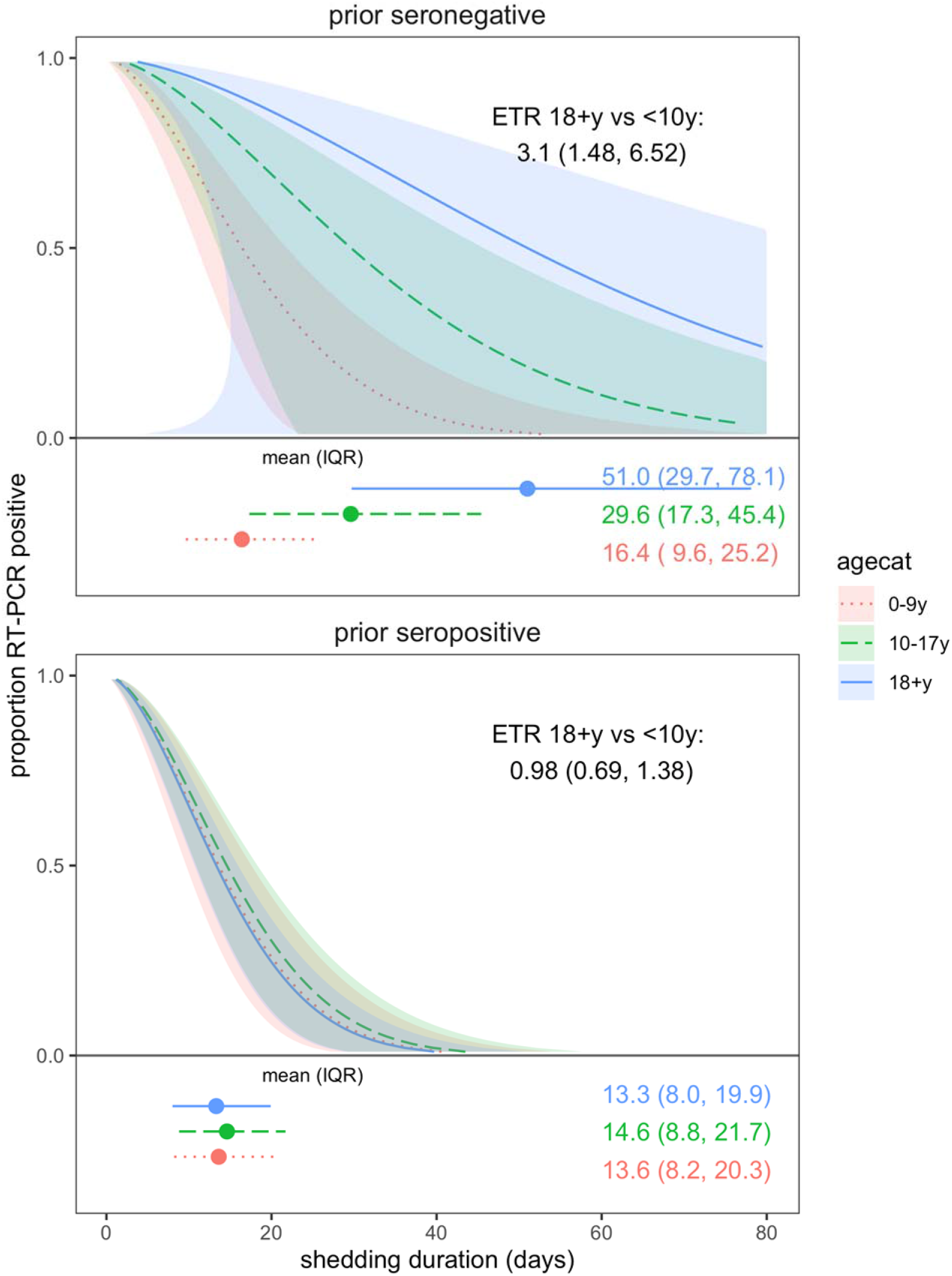
SARS-CoV-2 viral shedding duration by age among prior seronegative and seropositive. Results are from accelerated failure time (AFT) models. Shaded regions represent 95% confidence intervals. Individuals with ≥1 vaccination ≥14 days prior to infection were excluded.

## Notes

### Competing Interest Statement

Aubree Gordon serves on an advisory board for Janssen. All other authors report no competing interests.

### Author Declarations

This study was approved by the institutional review boards at the Nicaraguan Ministry of Health and the University of Michigan (HUM00119145 and HUM00178355). Informed consent or parental permission was obtained for all participants. Assent was obtained from children aged ?6 years.

## REFERENCES

Abu-Raddad, L. J., Chemaitelly, H., Ayoub, H. H., Tang, P., Coyle, P., Hasan, M. R., … Bertollini, R. (2022). Relative infectiousness of SARS-CoV-2 vaccine breakthrough infections, reinfections, and primary infections. Nat Commun, 13(1), 532. doi:10.1038/s41467-022-28199-7

Amanat, F., Stadlbauer, D., Strohmeier, S., Nguyen, T. H. O., Chromikova, V., McMahon, M., … Krammer, F. (2020). A serological assay to detect SARS-CoV-2 seroconversion in humans. Nat Med, 26(7), 1033–1036. doi:10.1038/s41591-020-0913-5

Cevik, M., Tate, M., Lloyd, O., Maraolo, A. E., Schafers, J., & Ho, A. (2021). SARS-CoV-2, SARS-CoV, and MERS-CoV viral load dynamics, duration of viral shedding, and infectiousness: a systematic review and meta-analysis. Lancet Microbe, 2(1), e13–e22. doi:10.1016/S2666-5247(20)30172-5

Chu, D. K. W., Pan, Y., Cheng, S. M. S., Hui, K. P. Y., Krishnan, P., Liu, Y., … Poon, L. L. M. (2020). Molecular Diagnosis of a Novel Coronavirus (2019-nCoV) Causing an Outbreak of Pneumonia. Clin Chem, 66(4), 549–555. doi:10.1093/clinchem/hvaa029

Covid-19 Cumulative Infection Collaborators. (2022). Estimating global, regional, and national daily and cumulative infections with SARS-CoV-2 through Nov 14, 2021: a statistical analysis. Lancet. doi:10.1016/S0140-6736(22)00484-6

Hodcroft, E., & Neher, R. (2022, 2022-04-07). Phylogenetic analysis of SARS-CoV-2 clusters in their international context - cluster 21K.Omicron. Retrieved from https://nextstrain.org/groups/neherlab/ncov/21K.Omicron

Hubeaux, S., & Rufibach, K. (2015). SurvRegCensCov: Weibull Regression for a Right-Censored Endpoint with Interval-Censored Covariate. R package version 1.4. Retrieved from https://CRAN.R-project.org/package=SurvRegCensCov

Killingley, B., Mann, A., Kalinova, M., Boyers, A., Goonawardane, N., Zhou, J., … Chiu, C. (2022). Safety, tolerability and viral kinetics during SARS-CoV-2 human challenge. Nature Portfolio, preprint. doi:10.21203/rs.3.rs-1121993/v1

Maier, H. E. (2022). SARS-CoV-2_shedding (Version 409886a). GitHub. Retrieved from https://github.com/hannahma/SARS-CoV-2_shedding

Maier, H. E., Balmaseda, A., Ojeda, S., Cerpas, C., Sanchez, N., Plazaola, M., … Gordon, A. (2021). An immune correlate of SARS-CoV-2 infection and severity of reinfections. medRxiv. doi:10.1101/2021.11.23.21266767

Maier, H. E., Kuan, G., Saborio, S., Bustos Carrillo, F. A., Plazaola, M., Barilla, C., … Gordon, A. (2021). Clinical spectrum of SARS-CoV-2 infection and protection from symptomatic re-infection. Clin Infect Dis. doi:10.1093/cid/ciab717

R Core Team. (2021). R: A Language and Environment for Statistical Computing. Retrieved from https://www.R-project.org/

Ritchie, H., Mathieu, E., Rodés-Guirao, L., Appel, C., Giattino, C., Ortiz-Ospina, E., … Roser, M. (2020). Coronavirus Pandemic (COVID-19). Our World in Data. Retrieved from https://ourworldindata.org/coronavirus

Singanayagam, A., Hakki, S., Dunning, J., Madon, K. J., Crone, M. A., Koycheva, A., … Investigators, A. S. (2022). Community transmission and viral load kinetics of the SARS-CoV-2 delta (B.1.617.2) variant in vaccinated and unvaccinated individuals in the UK: a prospective, longitudinal, cohort study. Lancet Infect Dis, 22(2), 183–195. doi:10.1016/S1473-3099(21)00648-4

Thernau, T. (2021). A Package for Survival Analysis in R. R package version 3.2-13. Retrieved from https://CRAN.R-project.org/package=survival

Wickham, H., Averick, M., Bryan, J., Chang, W., D’Agostino McGowan, L., François, R., … Yutani, H. (2019). Welcome to the tidyverse. Journal of Open Source Software, 4, 1686. doi:10.21105/joss.01686

